# Post-Coronary Artery Bypass Graft Complications; Potential Causes and Risk Factors

**DOI:** 10.1101/2022.12.29.22284005

**Authors:** Basheer Abdullah Marzoog, Karpunkin Oleg Alexandrovich, Tremasov Maxim Nikolaevich, Kostin Sergey Vladimirovich

## Abstract

**Background:** Cardiovascular disease (CVD) remains the leading cause of death worldwide despite the coexistence of the current COVID-19 pandemic. Current emergency management involves revascularization of the coronary arteries.

**Aims:** Retrospectively evaluating the association between the used number of shunts and postoperative complications. And to evaluate the potential pre/postoperative risk factors for complications development.

**Objectives:** Several complications are reported after coronary artery bypass graft (CABG) surgery, such as postoperative arrythmia and postoperative stroke. However, the risk factors for the development remain not elaborated.

**Material and methods:** A retrospective cohort study involved 290 patients for the period 2017-2021 treated surgically for ischemic heart disease. The surgery includes shunts of the internal thoracic arteries with the post-occlusion coronary arteries. The number of shunts is varied depending on the size and number of occluded arteries. According to the number of shunts, the patient may be operated with artificial circulation (CPB; cardiopulmonary bypass), or without working heart (OFF pump; without artificial circulation. For statistical analysis, used T test, one way ANOVA test, and Pearson correlation test by using Statistica program.

**Results:** The most frequently reported complication is postoperative hydrothorax, 28 (11.20%) patients. Patients with CABG and left ventricle aneurysm plastic surgery repair had a longer aortic cross-clamp time, t-value -2.20413, p <0.028306. Furthermore, patients with CABG have less ejection fraction, t-value 5.10667, p < 0.000001. Patients with post-CABG left ventricle (LV) aneurysm had less ejection fraction, t-value -2.01070, p <0.045292. Furthermore, patients with post-CABG LV aneurysm had a longer CPB time, t value -5.58113, p < 0.000000. Patients with post-CABG LV aneurysm had a longer aortic cross-clamp time, t-value -4.72802, p < 0.000004. Patients with postoperative hydrothorax are with low BMI and longer CPB and Aortic cross-clamp time, t-value -2.33929, p <0.020021; t-value 3.83233, P < 0.000157; t-value 2.71109, p < 0.007119, respectively. Subsequently, post-operative hydrothorax increases the intensive care unit (ICU) and total hospitalization days, t-value 5.80811, p < 0.000000; t-value 7.37431, p < 0.000000, respectively. Patients who have preoperative progressive angina have higher number of complications, t-value 2.108504, p < 0.035866. Post myocardial infarction myocardial sclerosis patients (PMIMS) have a higher number of complications, t-value 2.516784, p < 0.012396. No statistical correlation between number of the complications and the number of the shunts. Furthermore, there was no statistical association between the used number of internal thoracic arteries and the number of complications. A direct correlation between number of complications and age/CPB time/ aortic cross-clamp time/ICU hospitalization days/total hospitalization days, r= 0.138565, 0.204061, 0.162078, 0.487048, 0.408381; respectively.

**Conclusions:** Postoperative complication rate associated with the pre-existence of progressive angina and PMIMS. Elderly people undergoing CABG are at higher risk of psychosis, arrythmia, longer total and ICU hospitalization days, and stroke. Advanced age, longed CPB time, prolonged Aortic cross-clamp time, long ICU hospitalization days, and long total hospitalization days are risks for more frequent post CABG complications.

**Others:** The number of complications is not associated with the death and alive status of the patients or with the number of shunts.

## Introduction

Coronary artery graft surgery (CABG) performed on patients with ischemic heart disease to relieve the discrepancy between demand and supply of heart tissue. CABG surgery is performed in two versions, with stopped heart (on-pump) or working heart (off-pump) with cardiopulmonary bypass (CPB). CABG surgery involves revascularization of the coronary arteries through grafting of vessels from the saphenous vein, brachial vein, internal thoracic arteries, internal mammary arteries, and intercostal thoracic arteries [1]. The surgical approach to CABG is performed endoscopically or by thoracotomy. Recent surgeries are performed using the Da-Venice robot to minimize the risk of complications and improve prognosis by reducing the recovery period [2, 3].

Post CABG complications are frequent and seen in all types of CABG surgery, off-pump and on-pump. The remarkable advancement in the current CABG techniques reduced the complications frequency, but the underlying risk factors and pathophysiological mechanisms of development of each complication remain unclear. Depending on the type of surgery, the risk of development of complication is different. Several complications are reported after the postoperative period, such as postoperative arrythmia, post-operative psychosis, postoperative stroke, postoperative myocardial infarction, postoperative hydrothorax, re-sternotomy, postoperative dyspnea, and sternal wound infection [3, 4]. The risk factors for development of post CABG include diabetes mellites, arterial hypertension, dyspnea, progressive angina, and post myocardial infarction (MI) sclerosis, advanced age, long CPB time, prolonged Aortic cross-clamp time, long ICU hospitalization days, and long total hospitalization days [5, 6].

The study sought to clarify the role of number of used shunts in the complication’s development and role of number of the used internal thoracic arteries in the complication’s emergence after CABG surgery.

## Materials and methods

A retrospective cohort study analyzed data from surgically treated patients for the period 2017-2021 for various degrees of coronary artery disease. Data collected from Mordovian Republic Hospital for the past 5 years and retrospectively analyzed. The consent of the patients has been taken for scientific purposes to analyze and publish the results of the study. The sample includes 290 patients aged 41.000-80.000 years (mean; standard error 61.363; 0.397797). The study involves 46 (15.86%) female and male 244 (84.14%) males. In 2017, 33 (11.37931 %) patients performed coronary bypass surgery. In 2018, the number increased to 35 (12.06897 %) patients. In 2019, the number significantly increased to 61 (21.03448%) patients. In 2020, the highest number of bypass surgeries performed, 84 (28.96552 %) patients. In 2021, 77 (26.55172%) patients underwent coronary bypass surgery.

For statistical analysis, we used the T test, one- and the two-way ANOVA test, and Pearson correlation test using Statistica program. (StatSoft, Inc. (2011). STATISTICA (data analysis software system), version 10. www.statsoft.com.).

## Results

Body mass index (BMI) range from 16.360-45.350 Kg/m^2^ (mean; standard error: 45.350; 0.241918). Ejection fraction ranged from 29.000-77.000 ml/min (mean; standard error: 55.772; 0.438403). 278 (95.86207%) patients underwent on-pump CABG surgery and 12 (4.13793 %) patients underwent off-pump CABG surgery. The period of artificial circulation in surgeries with CPB ranged from 0.000-431.000 min (M; ±m: 431.000; 2.661081). The time of ischemia ranged from 0.000-147.000 min (M; ±m: 55.466; 1.341570). Postoperative hospitalization (hereafter, postoperative and post-CABG will be used interchangeably) in the intensive care unit ranged from 1.000 to 27.000 days (M; ±m: 2.821; 0.140122). The total hospitalization days (ICU hospitalization days plus recovery hospitalization days) after surgery ranged 7.000-50.000 days (M; ±m: 11.722; 0.292762).

By risk factor for ischemic heart disease; diabetes mellites, dyspnea, progressive angina, and postmyocardial infarction (MI) sclerosis. Out of 289 patients, 66 (22.75862 %) have diabetes mellites, and 223 (76.89655 %) without diabetes. Diabetes complications such as retinopathy and kidney injury reported in 28 (9.65517 %) patients and 261 (90.00000%) patients had no direct diabetes complications. Dyspnea exists in 55 (18.96552 %) patients and 234 (80.68966%) are negative. Various types of arrythmia present in 69 (23.79310 %) and 220 (75.86207 %) did not have arrythmia. Progressive angina reported in 112 (38.62069 %) patients and 177 (61.03448 %) don’t have progressive angina. Post-myocardial infarction sclerosis has been observed in 178 (61.37931 %) patients and 111 (38.27586 %) patients who did not have post-myocardial infarction sclerosis.

The preoperative left ventricle aneurysm present in 9 (3.10345 %) patients and 280 (96.55172 %) patients was negative. The number of patients who had shunt surgery and plastic surgery repair of the left ventricle is 7 (2.41379 %) patients and 283 (97.58621 %) patients who had only shunt surgery. The number of the patients who had one shunt is 30 (10.34483 %) patients, two shunts 67 (23.10345 %) patients, three shunts 119 (41.03448 %) patients, four shunts 65 (22.41379 %) patients, five shunts 8 (2.75862 %) patients, and six shunts 1(0.34483 %) patient. Depending on the number of shunts, the number of internal thoracic arteries (ITA) used. Patients who had one internal thoracic artery is 191 (65.86207 %) and 99 (34.13793%) patients had two internal thoracic arteries used in the shunt.

In terms of complications after shunt surgery, of 289 patients, 31 (10.73%) patients had postoperative arrythmia, 255 (88.24%) patients did not have postoperative arrythmia. Out of 283 patients, 14 (4.95%) patients had postoperative psychosis and 269 (95.05%) did not develop postoperative psychosis. A postoperative stroke developed in 4 (1.41%) patients and 279 (98.59%) were negative. Postoperative myocardial infarction occurred in 2 (0.71%) patients and 281 (99.29%) patients were negative. Postoperative hydrothorax reported in 34 (12.01%) patients and 249 (87.99%) patients didn’t report hydrothorax. Re-sternotomy registered in 9 (3.18%) patients and 274 (96.82%) patients didn’t have Re-sternotomy. Superficial postoperative sternal wound infection reported in 1 (0.40%) patient and 5 (2.00%) patients had deep postoperative sternal wound infection. Postoperative dyspnea reported in 11 (3.89%) patients and 272 (96.11%) patients didn’t have dyspnea.

Of 290 patients, 5 (1.72414%) passed (3 on the surgical table under anesthesia) and 285 (98.27586%) patients are alive. Out of 286 patients, 53 (18.27586 %) patients developed one complication (postoperative arrythmia, postoperative psychosis, postoperative stroke, postoperative myocardial infarction, postoperative hydrothorax, re-sternotomy, and postoperative dyspnea), two complications in 15 (5.17241%) patients, three complications in 3 (1.03448 %) patients, and four/five complication 1 (0.34483%) for each.

The most frequently reported complication is postoperative hydrothorax, 28 (11.20%) patients. By grouping the sample by gender, the BMI in women (mean 30.695 Kg/m^2^) is greater than in men (mean 28.444 Kg/m^2^), t-value -3.46865, p< 0.000603. Also, men (mean 60.630 years) are early affected by ischemic heart disease than women (mean 65.239 years), t-value -4.37030, p <0.000017. Furthermore, the ejection fraction of women (mean 57.978 ml) is higher than that of men (mean 55.354 ml), t-value -4.37030, p <0.000017. Progressive angina seen in individuals with high BMI (mean 29.406 Kg/m^2^), t-value -1.99512, p<0.046975. Diabetes mellites (DM) is seen in elderly patients (mean 63.288 years), t-value -2.65962, p <0.008262. Patients with DM are having higher BMI (mean 30.196 Kg/m^2^), t-value -3.18160, p < 0.001625. Also, patients with DM have longer inpatient hospitalization in the intensive care unit (ICU) and total hospitalization days, mean 3.580 days, t-value -3.02226, p <0.002739; mean 13.347, t-value - 3.10156, p <0.002120, respectively.

Moreover, DM complications are seen in obese people with high BMI (mean 30.600 Kg/m^2^), t-value -2.45502, p <0.014681. DM complications associated with longer ICU hospitalization days (5.011 days) and total hospitalization days (mean 16.854 days), t-value -5.41994, p <0.000000; t-value -6.16561, p <0.000000, respectively.

Shortness of breath results in an extension of total hospitalization days (mean 13.080 days), t-value 2.25940, p< 0.024627. The mean age of patients with preoperative arrythmia (mean 63.319 years) is higher than of patients without arrythmia (mean 60.776), t-value -2.75395, p <0.006265.

Post myocardial infarction myocardial sclerosis (PMIMS) statically reduces ejection fraction (mean 53.579 ml/beat) in compare with patients without post myocardial infarction myocardial sclerosis (mean 59.288 ml/beat), t-value 6.81607, p <0.000000. Furthermore, patients with PMIMS had a longer hospitalization in the ICU (mean 3.053 days) compared to patients without PMIMS (mean 2.450 days), t-value 0.035950, p <2.450. Patients with left ventricle aneurysm have significantly lower ejection fraction, t-value 4.85773, p <0.000002.

In terms of post-operative complications, patients with coronary artery bypass graft (CABG) and plastic surgery repair of the left ventricle aneurysm had longer Aortic cross-clamp time, t-value - 2.20413, p <0.028306. In addition, patients with CABG have less ejection fraction, t-value 5.10667, p < 0.000001. Patients with post-CABG left ventricle (LV) aneurysm had less ejection fraction, t-value -2.01070, p <0.045292. Additionally, patients with post-CABG LV aneurysm had a longer CPB time, t-value -5.58113, p < 0.000000. Also, Patients with post-CABG LV aneurysm had longer Aortic cross-clamp time, t-value -4.72802, p < 0.000004.

Post-CABG arrythmia developed in elderly patients, t-value -2.51307, p<0.012528. Subsequently, post-CABG arrythmia increased ICU and total hospitalization days, t-value - 5.83306, p< 0.000000; t-value -4.02907, p< 0.000072, respectively. Elderly people are at higher risk of psychosis after CABG surgery, t-value 2.69885, p <0.007379. Postoperative psychosis significantly increases the ICU hospitalization days, t-value 3.86094, p < 0.000140.

Postoperative stroke occurs more frequently in elderly people, t-value 2.087585, p < 0.037736. Subsequently, postoperative stroke increases the ICU hospitalization days, t-value 3.409293, p<0.000747.

Patients with postoperative hydrothorax have low BMI, t-value -2.33929, p <0.020021. Patients with postoperative hydrothorax had a longer CPB time, t-value 3.83233, P < 0.000157. Also, patients with postoperative hydrothorax had a longer Aortic cross-clamp time, t-value 2.71109, p < 0.007119. Subsequently, the postoperative hydrothorax increases the ICU and total hospitalization days, t-value 5.80811, p < 0.000000; t-value 7.37431, p < 0.000000, respectively. Patients with postoperative dyspnea stayed longer in the ICU and total hospitalization days, t-value 6.22816, p < 0.000000; t-value 7.17446, p < 0.000000, respectively. Superficial post-operative sternal wound infection increased ICU hospitalization days, t-value -3.66815, p < 0.000299. However, patients with deep postoperative sternal wound infection have longer ICU and total hospitalization days, t-value -2.61051, p < 0.009591; t-value -5.87749, p < 0.000000, respectively. Passed patients have a longer CPB and Aortic cross-clamp time, t-value -6.37465, p < 0.000000; t-value -2.46336, p < 0.014349, respectively. There are no statistical differences in the number of complications between living and deceased patients.

The number of ITAs used is less in the elderly people, t-value 2.41992, p < 0.016145. Patients with long Aortic cross-clamp time used two thoracic arteries, t-value -5.21934, p < 0.000000. Also, patients with two thoracic arteries had longer CPB time, t-value -3.91720, p < 0.000112. The Aortic cross-clamp time and the CPB time are in direct association with the number of shunts. (Figure 1)

**Figure 1:**
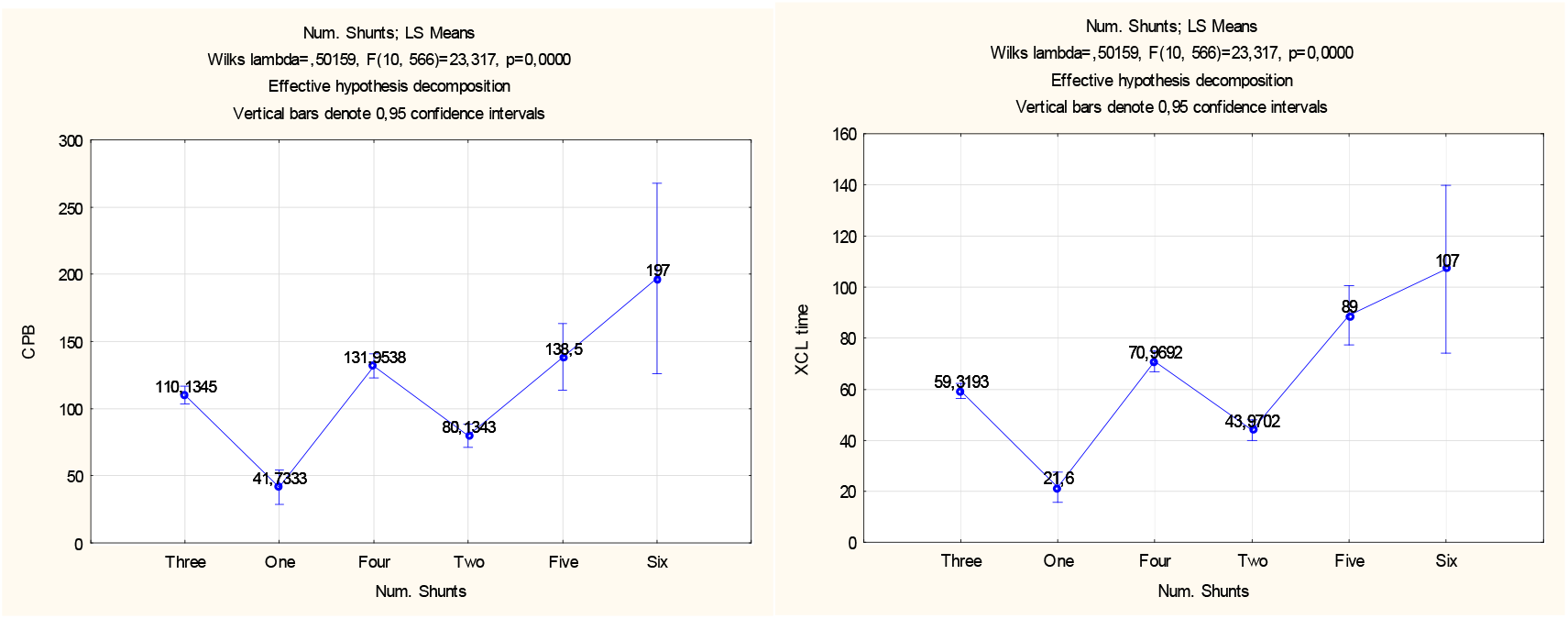
The Aortic cross-clamp time (XCL) and the CPB time are in direct association with the number of shunts.

Patients who have preoperative progressive angina have higher number of complications, t-value 2.108504, p < 0.035866. Patients with PMIMS have a higher number of complications, t-value 2.516784, p < 0.012396. No statistical significance in the correlation between number of the complications and the number of the shunts as well as between the number of used internal thoracic arteries and number of complications.

In terms of correlation, we found a direct association between age and ICU / total hospitalization days/number of complications, r= 0.189046, 0.141415, 0.138565; respectively. Furthermore, a direct correlation between the number of complications and age/CPB time/ Aortic cross-clamp time/ICU hospitalization days/total hospitalization days, r= 0.138565, 0.204061, 0.162078, 0.487048, 0.408381; respectively. A straight correlation between total hospitalization days and ejection fraction/ICU hospitalization days, r= 0.124180, 0.727443; respectively. The longer the Aortic cross-clamp time, the longer CPB period, r= 0.826151. A direct relation between BMI and ejection fraction, r= 0.119747. (Figure 2)

**Figure 2:**
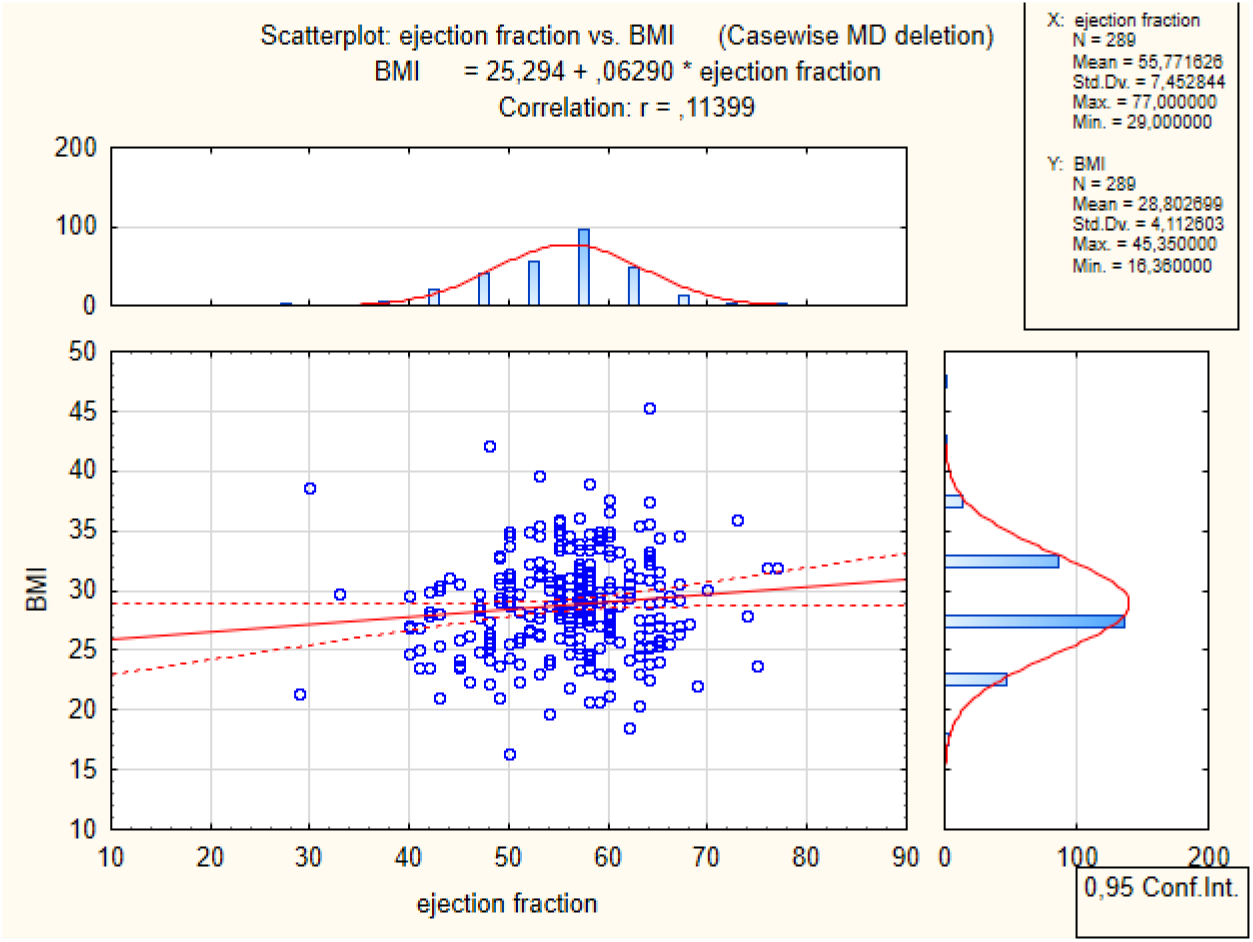
Direct proportional correlation between BMI and Ejection fraction.

## Discussion

In the light of results, prolonged Aortic cross-clamp time is seen in coronary artery bypass graft (CABG) and plastic surgery repair of the left ventricle aneurysm and patients with preoperative LV aneurysm. The prolonged Aortic cross-clamp time results in the use of two internal thoracic arteries. Prolonged aortic cross-clamp time increases hydrothorax risk and number of shunts which subsequently increases CPB time. Patients with low BMI are at high risk of hydrothorax. Furthermore, hydrothorax and the use of two internal thoracic arteries increase the time of the CPB. The coexistence of CABG and left ventricle aneurysm increases the time required for CPB. The post CABG ICU hospitalization days is related with the presence of post-operative arrythmia, post-operative psychosis, post-operative stroke, post-operative hydrothorax, post-operative dyspnea, and superficial sternal wound infection. Total hospitalization days depend on the presence of arrythmia, which is commonly seen in elderly patients > 63 years old. In addition, deep sternal wound infection increases total hospitalization days and ICU hospitalization days. The ejection fraction and the days of hospitalization in the ICU are closely associated with total days of hospitalization. Ejection fraction is age, sex, and presence of CABG and or CABG with LV aneurysm plastic repair dependent.

Age is a crucial factor in terms of postoperative complications. Where advance in age increases the ICU hospitalization days, total hospitalization days, and number of the complications. Elderly people are at high risk for stroke, psychosis, and arrythmia [4, 7–14]. However, elderly people passed one internal thoracic artery CABG.

The number of complications is not associated with the death and alive status of the patients or with the number of shunts. Furthermore, there was no statistical difference in the number of complications and number of the used shunts on number of the used internal thoracic arteries. However, the number of complications depends on the existence of preoperative progressive angina and PMIMS. In addition, the number of complications is determined by age increase, CPB time, the Aortic cross-clamp time, the days of hospitalization in the ICU, and total hospitalization days.

The results of our study are consistent with other studies. Several studies reported that post-CABG complications include arrythmia, particularly atrial fibrillation [4, 15–21]. However, the potential underlying pathophysiological mechanisms remain unclear and further elaboration is required. However, the potential pathophysiological pathway is multifactorial and involves autophagy dysfunction, upregulated sympathetic tone, mitochondrial dysfunction, inflammation, abnormal atrial conduction, and systemic inflammatory response indicated by elevated C-reactive protein and leukocytes [22–31]. Some studies showed that on-pump CABG surgery has higher risk of atrial fibrillation than off-pump [32–35].

## Conclusions

Elderly people undergoing CABG are at higher risk of psychosis, arrythmia, longer total and ICU hospitalization days, and stroke. Hydrothorax is seen in a low BMI and a long Aortic cross-clamp time which leads to longer CPB time. Progressive angina and PMIMS increase the risk of postoperative complications, particularly hydrothorax.

## Data Availability

All data produced in the present study are available upon reasonable request to the authors

## List of abbreviations

CVD: Cardiovascular disease
COVID-19: Corona virus infectious disease-19
CABG: coronary artery bypass graft
CPB: cardiopulmonary bypass
BMI: Body mass index
MI: myocardial infarction
DM: Diabetes mellites
ICU: intensive care unit
PMIMS: post myocardial infarction myocardial sclerosis
LV: left ventricle
ITA: internal thoracic artery

## Declarations

1. Ethics approval and consent to participate: applicable.
2. Consent for publication: applicable on reasonable request.
3. Availability of data and materials: The data are applicable.
4. Competing interests: The authors declare that they have no competing interests regarding the publication.
5. Funding: Not applicable (This research did not receive any specific grant from funding agencies in the public, commercial, or not-for-profit sectors).
6. Authors’ contributions: MB analyzed the statistical data, wrote the draft, and revised the final version of the paper, KS collected the data from the hospital. All authors have read and approved the manuscript.
7. Acknowledgments: none.
8. Authors’ information: Basheer Abdullah Marzoog, medical school student at National Research Mordovia State University (, +79969602820). Address: Bolshevitskaya st., 68, Saransk, Rep. Mordovia, 430005. ORCID: 0000-0001-5507-2413. Scopus ID: 57486338800.
9. The paper has not been submitted elsewhere.
10. The study approved by the National Research Mordovia State University, Russia, from “Ethics Committee Requirement N8/2 from 30.06.2021”. Written informed consent was obtained from the patients for publication of study results and any accompanying images.

